# Emergence of a novel reassortant Oropouche virus drives persistent human outbreaks in the Brazilian Amazon region from 2022 to 2024

**DOI:** 10.1101/2024.07.23.24310415

**Authors:** Felipe Gomes Naveca, Tatiana Amaral Pires de Almeida, Victor Souza, Valdinete Nascimento, Dejanane Silva, Fernanda Nascimento, Matilde Mejía, Yasmin Silva de Oliveira, Luisa Rocha, Natana Xavier, Janis Lopes, Rodrigo Maito, Cátia Meneses, Tatyana Amorim, Luciana Fé, Fernanda Sindeaux Camelo, Samyly Coutinho de Aguiar Silva, Alexsandro Xavier de Melo, Leíse Gomes Fernandes, Marco Aurélio Almeida de Oliveira, Ana Ruth Arcanjo, Guilherme Araújo, Walter André Júnior, Renata Lia Coragem de Carvalho, Rosiane Rodrigues, Stella Albuquerque, Cristiane Mattos, Ciciléia Silva, Aline Linhares, Taynã Rodrigues, Francy Mariscal, Márcia Andréa Morais, Mayra Marinho Presibella, Nelson Fernando Quallio Marques, Anne Paiva, Karina Ribeiro, Deusilene Vieira, Jackson Alves da Silva Queiroz, Ana Maísa Passos-Silva, Lígia Abdalla, João Hugo Santos, Regina Maria Pinto de Figueiredo, Ana Cecília Ribeiro Cruz, Livia Neves Casseb, Jannifer Oliveira Chiang, Livia Vinhal Frutuoso, Agata Rossi, Lucas Freitas, Túlio de Lima Campos, Gabriel Luz Wallau, Emerson Moreira, Roberto Dias Lins Neto, Laura W. Alexander, Yining Sun, Ana Maria Bispo de Filippis, Tiago Gräf, Ighor Arantes, Ana I. Bento, Edson Delatorre, Gonzalo Bello

## Abstract

The Brazilian western Amazon region is currently experiencing its largest laboratory confirmed Oropouche virus (OROV) outbreak, with nearly 6,000 reported cases in the states of Amazonas (AM), Acre (AC), Rondônia (RO), and Roraima (RR), between August 2022 and March 2024. Here, we sequenced and analyzed 382 full-length OROV genomes from human samples collected between 2022 and 2024 from all four states, aiming to trace the origin and genetic evolution of OROV leading to the current outbreak. Genomic analyses revealed that the recent upsurge of OROV cases in the Brazilian Amazon region coincides with the emergence of a novel reassortant viral lineage containing the M segment of viruses detected in the eastern Amazon region from 2009 to 2018 and the L and S segments of viruses detected in Peru, Colombia, and Ecuador from 2008 to 2021. The novel reassortant OROV lineage likely emerged in the Central region of the AM state between 2010 and 2014 and displayed a long-range silent dispersion during the second half of the 2010s. The 2022-2024 OROV epidemic was spatially segregated into three major subpopulations located in RR, AMACRO (a bordering region between AC, RO, and AM-Southern region), and AM-Central (which includes the Amazonas’ capital, Manaus) regions. The peak of OROV transmissions in all regions occurred during the rainy season in the Amazon basin. Furthermore, our phylodynamics reconstructions showed that OROV spread was driven mainly by short-range (< 2 km) movements, with an average dispersal rate ≤ 1.2 km/day, consistent with the pattern of an active flight of infected vectors. Nevertheless, a substantial proportion (22%) of long-range (> 10 km) OROV migrations were also detected, consistent with viral dispersion via human activities. Our data provides an unprecedented view of the real-time spread and evolution of a neglected emergent human pathogen. Moreover, our results emphasize the need for widespread, long-term genomic surveillance to better understand the real burden of OROV within and beyond the Amazon region.

## Main Text

Oropouche virus (OROV) is an arthropod-borne pathogen of the family *Peribunyaviridae,* genus *Orthobunyavirus,* species *Orthobunyavirus oropoucheense.* OROV is an endemic zoonotic arbovirus in the Amazon region that is maintained in a sylvatic transmission cycle among vertebrate hosts such as sloths, rodents, non-human primates, and birds by different vector species, including *Aedes serratus, Psorophora cingulata,* and *Haemagogus tropicalis*.^1,2^ Similarly to the yellow fever virus (YFV), and in contrast to urban arboviruses such as dengue (DENV), Zika (ZIKV), and chikungunya (CHIKV), human infections are not essential to maintain the OROV life cycle. However, occasional OROV spillovers from wildlife to humans can initiate localized outbreaks or large epidemics once the virus is introduced to rural or urban settings^1^. OROV transmission in anthropogenic environments seems to be driven mainly by the biting midges *Culicoides paraensis*, although *Culex quinquefasciatus* mosquitoes have also been suggested as a possible secondary vector.^1,3–5^ At least 30 human outbreaks of OROV have been irregularly reported in the Amazon region since the 1960s.^6^

Until the emergence of CHIKV and ZIKV in 2014-2015, OROV was the arbovirus with the second highest incidence in Brazil, only surpassed by DENV.^7^ The country’s largest documented OROV outbreak was reported in the Pará (PA) state in the late 1970s, where estimates based on serological, clinical, and epidemiology data pointed to >100,000 human cases ^8^. During the 21^st^ century, small outbreaks of OROV were documented in Manaus, the capital city of the Amazonas (AM) state in 2007-2008,^9^ in a rural community of the Amapá (AP) state in 2008-2009^10^, and in rural settings of the PA state in 2003-2004, 2006 and 2018.^11–13^ However, the burden of OROV infections in Brazil has probably been underestimated due to the lack of systematic laboratory differential diagnosis and clinical symptoms similar to those of other endemic arboviral diseases.^14^ Moreover, sporadic cases of OROV infection were detected between 2004 and 2016 in several municipalities of the AM, Acre (AC), Bahia, Mato Grosso, and Mato Grosso do Sul states with no reported outbreaks of this viral disease.^3,14–20^ These data support a silent transmission and expansion of the OROV geographical boundaries within Brazil.

Between late 2022 and early 2024, the Brazilian states of AC, AM, Rondônia (RO), and Roraima (RR) located in the western Amazon region reported a sharp increase in the incidence of OROV cases.^21^ As of 31^st^ March 2024, 6,226 have tested positive for OROV (OROV+) by a real-time RT-PCR protocol^22^ in both rural areas and major urban centers of the western Amazon region (**Fig. 1a**). AM reported the highest number, with 3,685 notified cases, followed by RO (1,756), AC (441), and RR (344). This OROV epidemic is not only the most significant upsurge of human OROV cases in the 21^st^ century but also the most geographically spread OROV epidemic ever documented, affecting over 140 municipalities from the Amazon basin (**Fig. 1a**), highlighting the risk of viral exportation and community transmission outside the Amazon basin. Since early 2024, dozens of autochthonous cases of OROV have been reported in non-Amazonian Brazilian states.^23,24^ Thus, there is a crucial need to enhance our comprehension of the underlying mechanisms that fueled the current transmission of OROV in the Amazon region.

**Figure 1.**
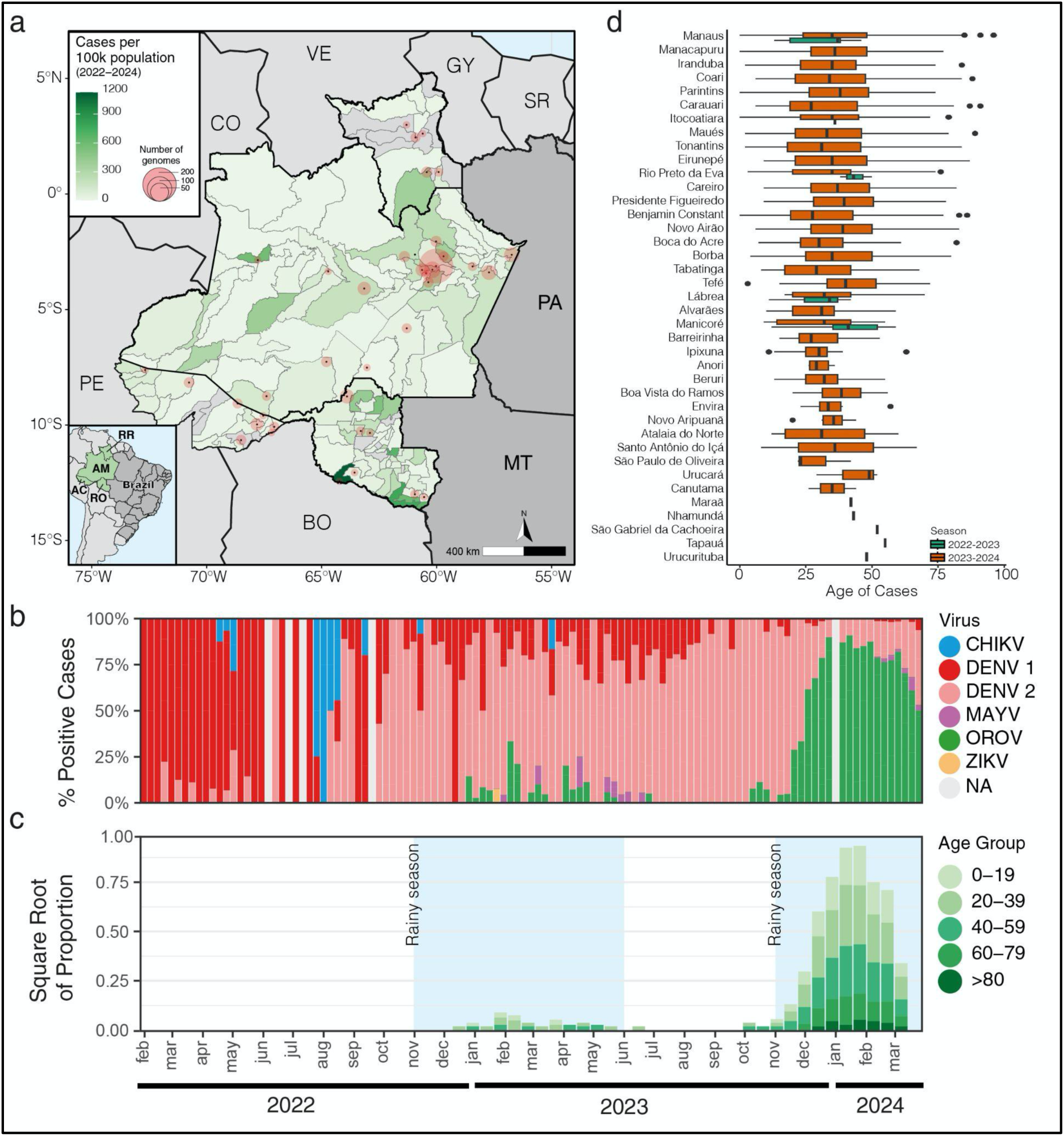
Epidemiological characteristics of the OROV outbreak in the western Amazonian Brazilian region (2022-2024). **a**) Geographic distribution of the OROV+ cases per 100,000 population diagnosed in Amazonas (AM), Acre (AC), Roraima (RR), and Rondônia (RO) between January 2022 and April 2024. Each municipality is colored based on the number of confirmed OROV-positive cases, as indicated by the color gradient in the top left legend. Additionally, circles represent the locations where OROV genomes were obtained, with circle sizes proportional to the number of genomes generated, as shown in the legend. The map also includes neighboring Brazilian states Pará (PA) and Mato Grosso (MT); as well as neighboring countries: Bolivia (BO), Peru (PE), Colombia (CO), Venezuela (VE), Guyana (GY), and Suriname (SR). **b**) Molecular positivity for arboviruses in the AM state, Brazil, from February 2022 to March 2024. **c**) Square root of the proportion of OROV-diagnosed cases biweekly, separated by age group. Blue-shaded areas indicate the Amazon basin region’s rainy season (November through May). **d**) Box plot of the age distribution of OROV cases by municipality, illustrating the median and interquartile range (IQR) for the seasons 2022-2023 (green) and 2023-2024 (red).

We then inspected the temporal pattern of OROV+ cases in the AM state, where extensive OROV testing occurred between November 2022 and March 2024 (**Fig S1a**). The data revealed that a very small first wave of OROV cases was recorded between late 2022 and early 2023, followed by a more significant second wave starting in October 2023, which dominated the epidemiological landscape of arboviral infections from December 2023 to March 2024 (**Fig. 1b**). The peak of OROV+ cases in the second wave coincides with the rainy season (from November to April) in the Amazon region (**Fig. 1c**), in line with previous OROV outbreaks/epidemics described in Brazil.^25^ Rainfall after the dry season replenishes the soil with moist sites suitable for oviposition, immature development, and emergence of adult midges and coincides with higher levels of vector abundance.^26^ Increased rainfall was associated with an expansion of the *C. paraensis* population in different settings of northern Brazil and in northwestern Argentina.^27^ It was estimated that there would be a lag time of approximately 1-2 months between the initial increase in rainfall and the corresponding increase in the midge population in the Amazon biome.^28^ These findings underscore the significant indirect effect of pluviosity on the increased transmission of OROV in the Amazon region, probably through its influence on the lifecycle of sylvatic/urban vectors.

The sylvatic nature of OROV transmission raises the hypothesis that potential vectors predominantly remain outside households, thereby increasing exposure risks for young, working-age males employed in forestry or other activities with frequent forest-dwelling animals/vector interaction. However, our results indicate that individuals from all age groups were infected with OROV (**Fig. 1c**). Additionally, the age distribution of infections was consistent across municipalities of varying sizes and degrees of urbanization (**Fig. 1d**). The adjustment of age distributions to a Gaussian model indicated a slight skew towards older ages among individuals diagnosed with OROV (35.7 years, SD = 17.0) compared to those with DENV (32.3 years, SD = 17.9 years) (**Fig. S1b**) and sex distribution indicates a roughly equilibrium between sexes, with a male to female ratio of 1.09:1 in OROV cases, comparable to that observed in DENV cases (1.02:1, *P* = 0.595) (**Fig. S1c**). Interestingly, we detected co-infections of OROV with Mayaro virus (MAYV, n=1), with DENV type 1 (n=1) and with DENV type 2 (n=8) (**Fig S1d**). These findings suggest that the transmission of OROV follows a pattern like other urban arboviruses and highlights the need for integrated vector control strategies targeting both mosquitos and midges in urban environments of the Amazon region. Viral co-infections, albeit at low frequencies, also underscore the complex epidemiological landscape of arboviral infections and the potential interactions between different arboviruses during concurrent outbreaks in urban settings.

Fever (97.2%), headache (92.7%), and myalgia (78.2%) were the symptoms more frequently reported among the 2,272 OROV-positive patients from the AM state who answered the medical form in 2024. Other reported symptoms included nausea (42.1%), retro-orbital pain (34.1%), arthralgia (28.5%), vomiting (25.7%), photophobia (8.4%), diarrhea (7.4%), rash (2.8%), and edema (2.8%). Interestingly, two of these symptoms were more frequently observed in female patients: nausea (*P* < 0.0001) and vomiting (*P* = 0.0005) (**Supplementary Table 1**). So far, no fatal cases have been attributed to the disease. Clinical manifestations in the current outbreak were consistent with those observed in previous epidemics, where the most reported symptoms were fever (73-100%), headache (73-94%), myalgia (64-87%), and arthralgia (58-85%). These were followed by chills (10-64%), photophobia (17-58%), retroocular pain (14-65%), dizziness (10-42%), nausea (24-36%), vomiting (24%), diarrhea (10-13%), and rash (2-42%).^9,11–13,29–35^ Of note, a significant proportion of patients (16%) infected in a previous OROV epidemic in Manaus in 2007-2008 had spontaneous hemorrhagic phenomena (petechiae, epistaxis, and gingival bleeding),^9^ that were not observed in our study population. Thus, most OROV infections during the 2023-2024 epidemic in the AM state were mild and self-limited, with similar clinical manifestations to previous epidemics, as was recently described in the RO state.^36^ These findings also support the notion that OROV diagnosis based solely on clinical symptoms is extremely difficult and reinforced the importance of implementation of accurate molecular diagnostic tests.^32^

To understand whether the 2022-2024 west Amazon Brazilian OROV epidemic was fueled by one or multiple zoonotic events, and to estimate when and where the virus entered human populations and how the virus changed and spread as the current epidemic unfolds, we sequenced full-length OROV genomes from 382 patients living in 37 municipalities of the Brazilian states AM (n = 321), AC (n = 27), RO (n = 22), and RR (n = 12), collected between 30^th^ August 2022 and 25^th^ February 2024 (**Fig. 1a**). These 382 sequences were combined with all full-length OROV genome sequences available at the NCBI (n = 72), sampled in the Americas between 1955 and 2021, including the prototype sequences of OROV (OROV_P_), Iquito virus (IQTV_P_), Perdoes virus (PEDV_P_), and Madre de Dios virus (MDDV_P_). Bayesian phylogenetic analyses revealed two large well-supported (*PP* ≥ 0.90) viral lineages for the M (M_1_ and M_2_) and L (L_1_ and L_2_) segments (**Fig. 2a-b**). The tree topology of the S segment was less resolved and sequences were dispersed across multiple clades (**Fig. 2c**), consistent with the notion that this segment contains less phylogenetic information than the segments M and L.^10,30,37^ Using a minimum cut-off of clade size (n > 10 sequences) and support (*PP* ≥ 0.90), we identified three major lineages (S_1_, S_2,_ and S_3_) and several minor lineages (S_X_) for the S segment (**Fig. 2c**). These observations support the relevance of developing a phylogenetic classification system based on all the OROV gene segments as previously established for other segmented viruses such as rotavirus.^38^ According to this system and the clades previously described, the OROV_P_ prototype is classified as M_1_L_1_S_1_, the IQTV_P_ as M_X_L_2_S_X_, the PEDV_P_ as M_X_L_2_S_3_, and the MDDV_P_ as M_X_L_X_S_X_.

**Figure 2.**
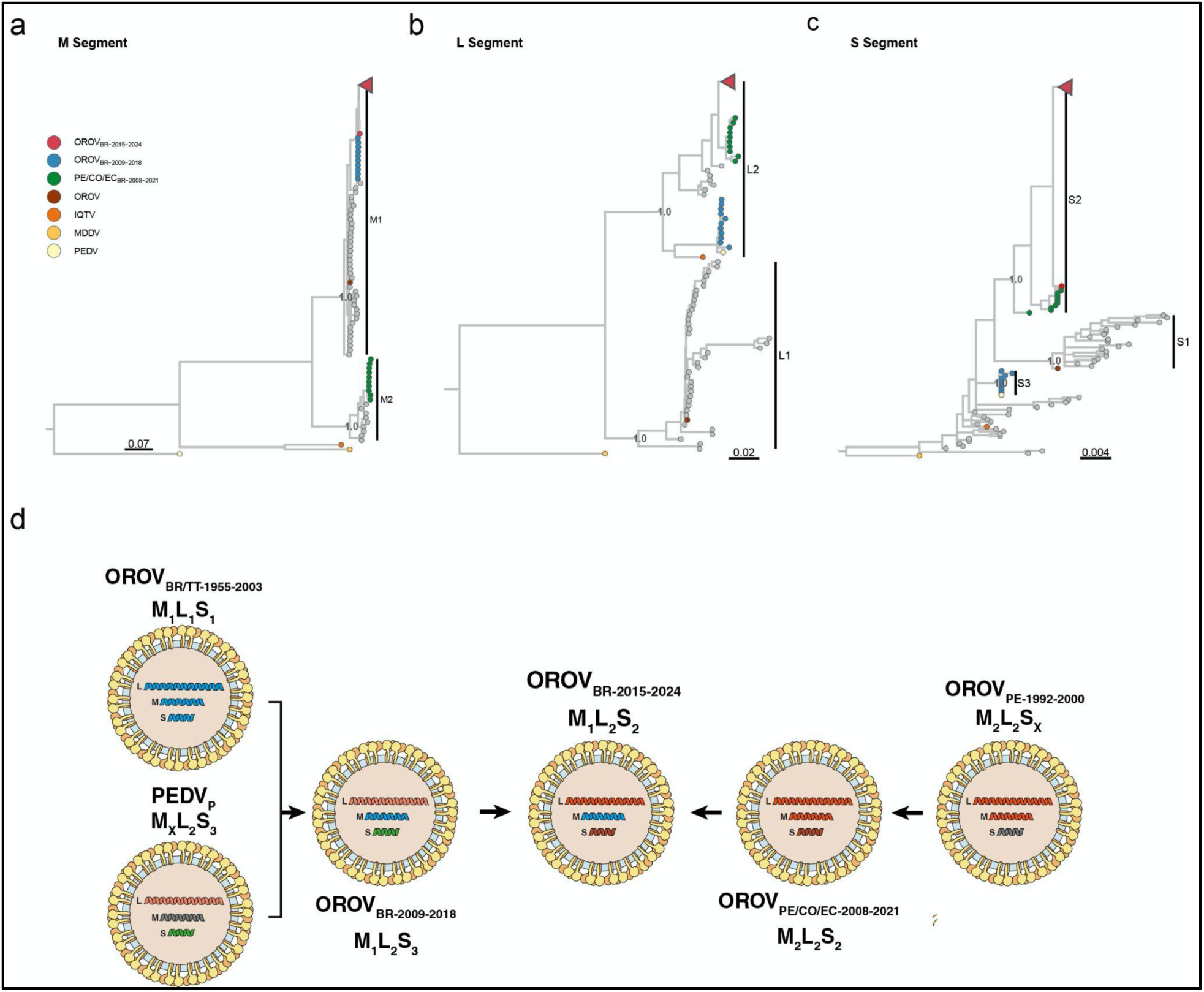
Phylogenetic analyses of the M, L, and S segments of OROV. **a-c)** Bayesian consensus tree inferred from the segments M (**a**), L (**b**), and S (**c**) of OROV sequences with complete genomes (n = 454). The tips of prototypical viruses and major clusters are color-coded according to the legend in the left corner. To improve visualization, the largest fraction of the OROV_BR2015-2024_ was collapsed and indicated in the figure by a red triangle. Brackets demarcate major monophyletic lineages alongside their denomination and posterior statistical support. All trees are drawn according to the genetic distance scale at the bottom of each panel. d) Putative reassortment events that generated the current genomic diversity of OROV in South America. Each OROV genomic segment is colored according to the lineage identified in this study. Illustrations were obtained from SwissBioPics (https://viralzone.expasy.org/,^44^) and modified to align with the reassortment events hypothesis. OROV: Oropouche virus, IQTV: Iquitos virus, MDDV: Madre de Dios virus, PEDV: Perdões virus.

The OROV sequences sampled in Brazil between 2022 and 2024 branched in a highly supported (*PP* > 0.90) monophyletic clade (OROV_BR-2015-2024_) across all three genomic segments together with a sequence sampled in French Guiana in 2020 ^31^ and another sequence sampled in Tefé, AM, Brazil, in 2015 ^14^ (**Fig. 2a-c**). The OROV sequence from Tefé-2015 branched as the most basal strain of the OROV_BR-2015-2024_ clade in all segments. The OROV sequence most closely related to the OROV_BR-2015-2024_ clade differed between the M and the L+S segments. The OROV_BR-2015-2024_ clade branched within the M_1_ lineage, which comprises all OROV sequences sampled in Brazil since 1960 and was most closely related to a sister monophyletic clade (OROV_BR-2009-2018_) that comprise sequences sampled in the eastern Amazonian states of AP in 2009 and PA in 2018 (**Fig. 2a**). In the L and S segments, the OROV_BR-2015-2024_ clade branched within the L_2_ and S_2_ lineages, nested within a basal paraphyletic clade (OROV_PE/CO/EC-2008-2021_) that encompass sequences sampled in Peru, Colombia, and Ecuador between 2008 and 2021 (**Figs. 2b-c**).

Interestingly, the OROV_BR-2009-2018_ clade branched with high support with the PEDV_P_ prototype in both L (L_2_ lineage) and S (S_3_ lineage) segments. Meanwhile, the OROV_PE/CO/EC-2008-2021_ clade branched within the M_2_ lineage, with sequences detected in Panama and Peru between 1989 and 2000 (**Fig. 2a-c**).

The considerable topological discordance among the phylogenetic trees of different genomic segments supports the occurrence of successive reassortment events throughout OROV evolution in South America, as schematically depicted in **Fig. 2d**. According to the proposed model, sequences sampled in Brazil between the 1960s and the 1990s mainly belong to the M_1_L_1_S_1_ lineage. In contrast, sequences sampled in Peru between 1992 and 2000 belong to the M_2_L_2_S_X_ lineage. The OROV_BR-2009-2018_ clade detected on the eastern Amazon between 2009 and 2018^13,39^ was probably a M_1_L_2_S_3_ reassortant virus that possesses the M_1_ segment from viruses already circulating in the Amazon region and the L_2_+S_3_ segments detected in the PEDV_P_ isolated from a non-human primate (NHP) outside the Amazon basin in 2012.^13,39^ The OROV_PE/CO/EC-2008-2021_ clade detected in Peru, Colombia, and Ecuador from 2008 to 2021^37^ was an M_2_L_2_S_2_ virus that combined the M_2_+L_2_ segments of viruses circulating in Peru during the 1990s, along with a new S_2_ segment that may have evolved from a basal S_X_ segment or may have been acquired from some unsampled OROV lineage. Finally, the OROV_BR-2015-2024_ clade was a M_1_L_2_S_2_ reassortant virus that acquired the M_1_ segment from the OROV_BR-2009-2018_ clade and the L_2_+S_2_ segments from the OROV_PE/CO/EC-2008-2021_ clade. Reassortment is a powerful mechanism driving the evolution of bunyaviruses, and consistent with our findings, most recognized reassortants orthobunyaviruses possess L+S segments derived from one virus and M segment from another.^10,40–43^ The balance between superinfection resistance, which promotes reassortments between distantly related viruses, and the compatibility of newly mixed components, which favors reassortments between closely related viruses, shapes productive exchanges.^43^

To track the time of the most recent common ancestor (T_MRCA_) of the OROV_BR-2015-2024_ clade and the time interval of the reassortment event, we reconstruct a Bayesian time-scale phylogenetic tree from the OROV_BR-2015-2024_ clade plus their most closely related sequences belonging to the OROV_PE/CO/EC-2008-2021_ clade in the L and S segments and the OROV_BR-2009-2018_ clade in the M segment. The correlation between genetic divergence and sampling time was significant for datasets of all three genomic segments (*p* < 0.05). However, the temporal structure of the M segment (Correlation coefficient = 0.94) was more pronounced than that of the L and S segments (Correlation coefficient ∼ 0.80) (**Fig. 3a-c**). The median estimated evolutionary rate for all three segments was roughly similar and ranged between 1.3 - 1.7 × 10^-3^ substitutions/site/year (**Fig. 3d**), although the shorter segment S presented higher estimates of uncertainty. Analyses of all three genomic segments traced back the median T_MRCA_ of the OROV_BR-2015-2024_ clade to 2013-2014, slightly before the sampling time of the oldest sequence from that clade detected in Tefé-AM in 2015. The time frame of the reassortment event between clades OROV_BR-2009-2018_ and OROV_PE/CO/EC-2008-2021_ was estimated between 2009 and 2013 (**Fig. 3e-g**). OROV reassortment events were also reconstructed using the CoalRe model (**Figure S2**). Due to the considerably large genetic diversity of the virus and multiple reassortment events that likely took place in the OROV lineage’s evolutionary history, MCMC for the CoalRe model only converged when a subsampled dataset of recent sequences (sampled from 2000 onward) was analyzed. The model estimated a reassortment rate of 0.1 [0.05 - 0.18, 95% highest posterior density (HPD)] events per year, which is very similar to what was previously estimated for p1918−like H1N1 and influenza B viruses.^45^ However, the posterior probability (*PP*) of most of the reassortment events reconstructed by CoalRe was very low (*PP* <10), and only segment M reassortments showed intermediate to high support (purple dashed lines in **Figure S2**). The reassortment event that originated the OROV_BR-2015-2024_ clade was highly supported (*PP* > 90) and probably occurred around 2010, in agreement with the pattern revealed by the independent analyses of the three viral segments. These findings support a short-term silent dispersion of the OROV_BR-2015-2024_ clade during the first half of the 2010s before being detected for the first time in the small inner city of Tefé in the AM state in 2015 and subsequently in French Guiana in 2020.^14,31^

**Figure 3.**
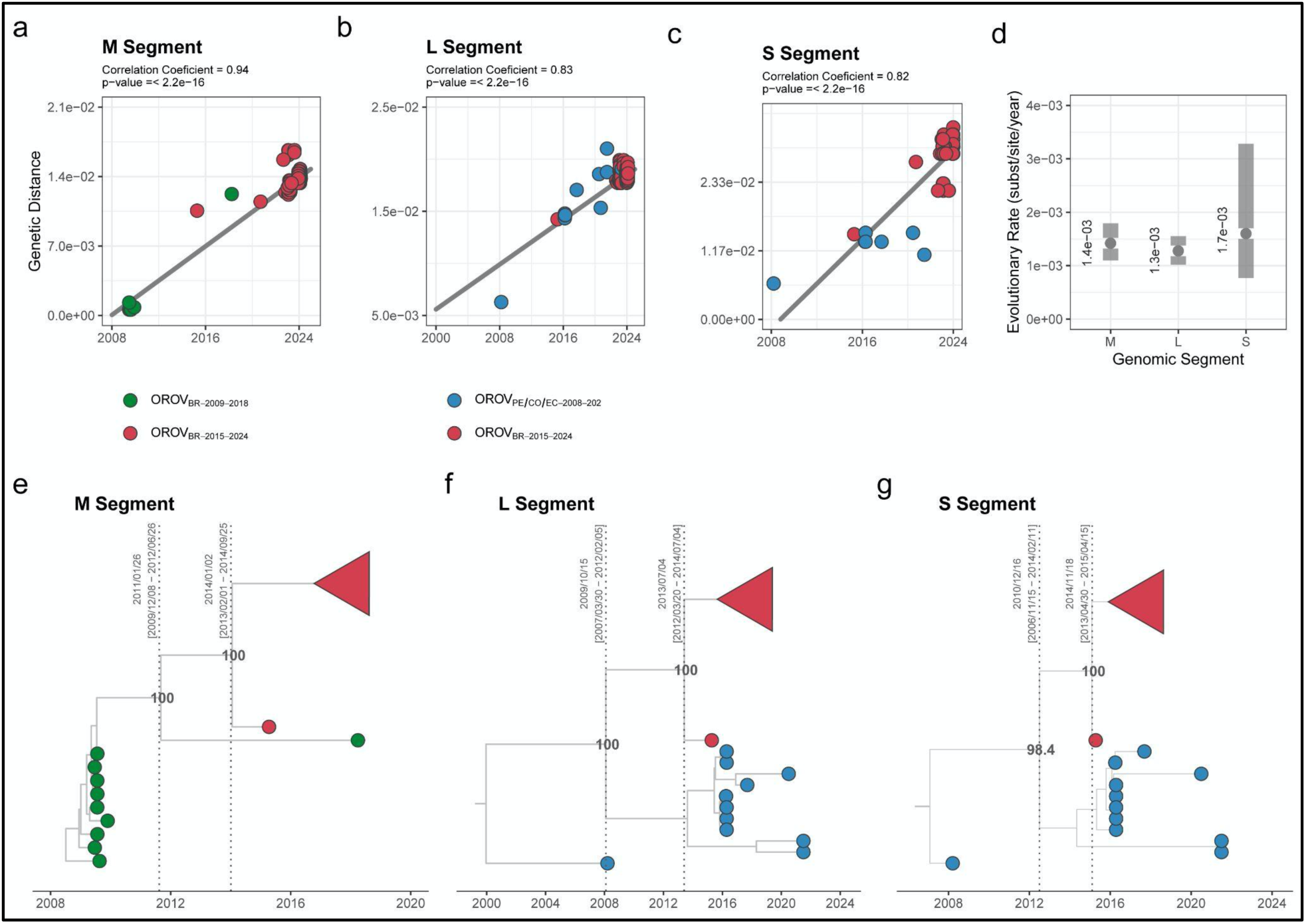
Temporal dynamics of the current outbreak OROV_BR-2015-2024_ clade and the most closely related clades. The figure represents the root-to-tip linear regression between collection date and genetic distance (**a-c**), molecular clock median value and 95% HPD (**d**), and time-scaled Bayesian maximum clade credibility tree (MCCT) (**e-g**) inferred with datasets composed of the OROV_BR-2015-2024_ clade (red tips) and its most closely related sequences of the OROV_BR-2009-2018_ clade (green tips) in the M segment (*n* = 87) and of the OROV_PE/CO/EC-2008-2021_ clade (blue tips) in the L (*n* = 88) and S (*n* = 88) segments. The posterior probability of key nodes in the MCCT are annotated. To improve visualization, the largest fraction of the OROV_BR-2015-2024_ was collapsed in the MCCT in all three segments, as indicated by a red triangle.

We used a Bayesian phylogeographic framework to reconstruct the spatial dissemination of the OROV_BR-2015-2024_ clade in the Amazon basin, analyzing a concatenated full-length genomic dataset. The genome-wide molecular clock rate of the OROV_BR-2015-2024_ clade was estimated at 1.2 × 10^-3^ (95% HPD: 1.1 - 1.4 × 10^-3^) substitutions/site/year. A discrete phylogeographic model was first employed to reconstruct the migration between states and regions in western Amazon: AC, RO, RR, Manaus, AM-Central, AM-South, AM-North, and AM-Southwest. Discrete phylogeographic reconstructions indicate that the OROV_BR-2015-2024_ clade was spatially and temporally segregated in six major highly supported (*PP* > 0.90) lineages (**Fig. 4a-b**). The monophyletic sub-clade RR-I was exclusively composed of sequences from RR from August 2022 to May 2023. The paraphyletic sub-clade AMACRO-I was exclusively composed of sequences sampled in AMACRO, a bordering region of 454,000 km² between AC, RO, and AM Southern region (**Fig. 4c**) between December 2022 and May 2023. The monophyletic sub-clade AMACRO-II comprises all OROV sequences detected in AC, RO, and AM-South region from December 2023 onwards. OROV sequences detected in Manaus and AM-Central/Northern regions from November 2023 to February 2024 branched within the monophyletic sub-clades AM-I (83%), AM-II (11%) and AM-III (6%). The OROV_BR-2015-2024_ clade was initially spread from the AM-Central region following two main routes (**Fig. 4a**). In the northward route, the virus reached RR (*PSP* = 1.0) around June 2022 and originated the RR-I sub-clade that drove multiple outbreaks in this state during the 2022-2023 rainy season, but we found no evidence of further dissemination beyond that period (**Fig. 4d**). In the southward route, the virus most probably first reached RO (*PSP* = 1.0) in August 2022 (**Fig. 4d**). However, this finding should be interpreted with caution due to uneven geographic sampling at late 2022, originating the AMACRO-I sub-clade. From RO, the AMACRO-I sub-clade rapidly spread to AC and AM-South, causing an epidemic wave in different municipalities of the AMACRO region during the 2022-2023 rainy season. This sub-clade persisted throughout the 2023 dry season and gave rise to four new sub-clades detected in the 2023-2024 rainy season: the AMACRO-II sub-clade most probably emerged in AC (*PSP* = 0.97) around August 2023 (**Fig. 4a-d**) and spread to RO and AM-South region, and the AM-I, AM-II and AM-III sub-clades that emerged in the city of Manaus (*PSP* > 0.95) between mid-August and early September 2023 (**Fig. 4a-d**). Subsequently, the AM-I and AM-II sub-clades spread from Manaus to other municipalities in the Central and Northern regions of AM state from December 2023 onwards (**Fig. S3**).

**Figure 4.**
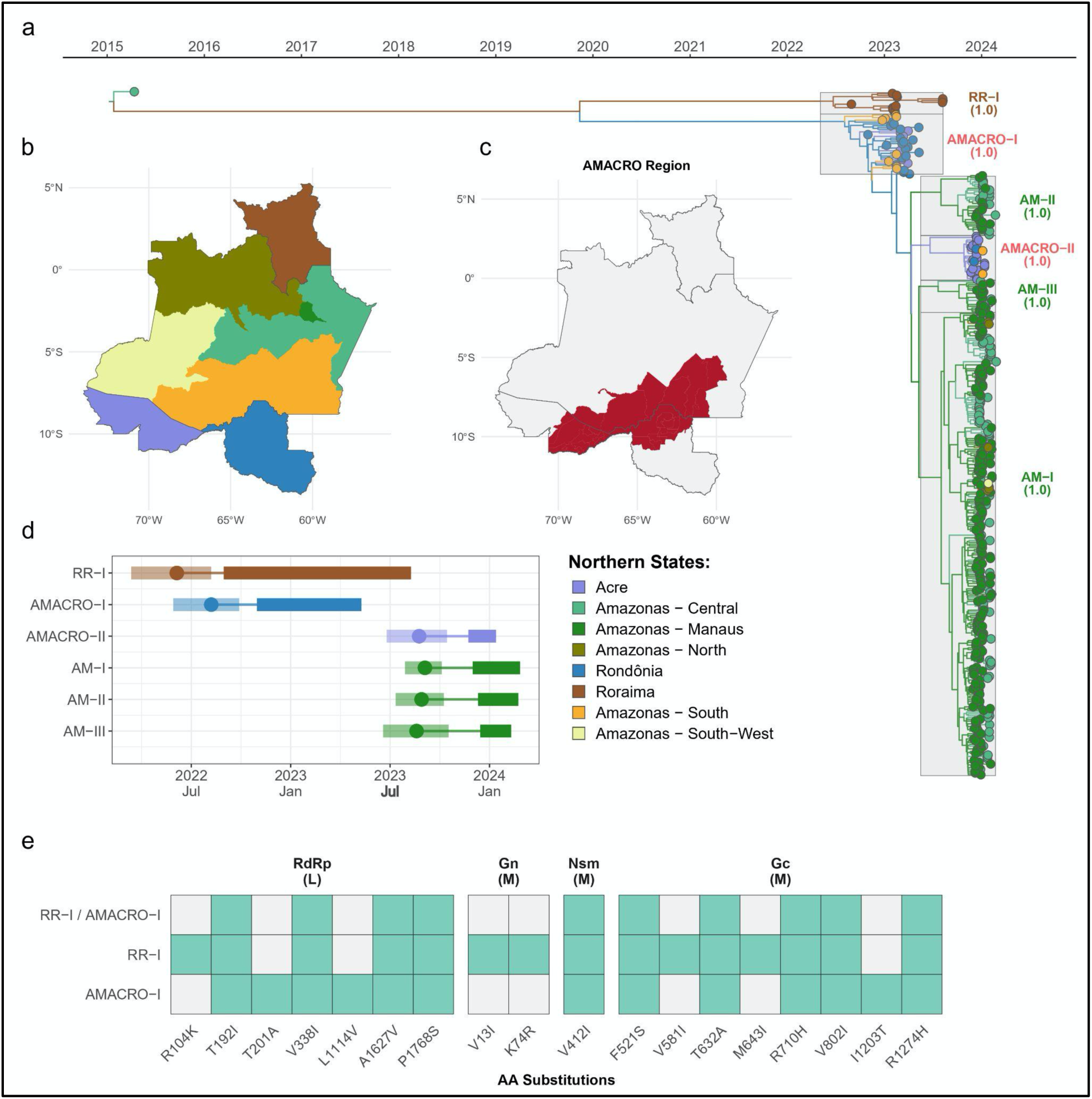
Spatial dissemination of the OROV_BR-2015-2024_ clade in the western Brazilian Amazon region. **a**) Bayesian time-scaled MCC tree of the OROV_BR-2015-2024_ clade, inferred after concatenation of the three genomic segments (L, M, and S) (*n* = 383). Branches are colored according to the inferred location of their ancestor nodes, and tips are colored according to sampling locations, both following the color scale shown in the bottom left corner of the tree. Sub-clades are highlighted in the tree alongside their posterior probabilities. **b-c**) Maps of the western Brazilian region, showing its constituent states, along with sub-state regions of Amazonas (**b**), both according to the same color scale used in the MCC tree and the supra-state AMACRO region (**c**). **d**) Temporal dynamics of major OROV_BR-2015-2024_ sub-clades. For each sub-clade, we represent the time of the most recent common ancestor (T_MRCA_, *circle*) and its 95% highest posterior density (HPD) interval (*transparent polygon*), the period of cryptic circulation (*thinner line*), and the sampling range (*thicker line*). Each sub-clade is colored following the same color scale used in the MCC tree. **e**) Amino Acid substitutions identified in the ancestral sequences of selected OROV clusters.

The phylogeographic analysis revealed that OROV sub-clades detected in AC, RO and AM in 2024 evolved from a single common ancestor, suggesting that a small population of closely related viruses persisted in the AMACRO region throughout the 2023 dry seasons and fueled epidemics in the 2023-2024 rainy season. To confirm this hypothesis, we reconstructed the viral demographic dynamic using a BSKG model. This analysis supports an expansion of the *Ne* from November to March in the RR/AMACRO regions and a significant decrease between April and October, indicating a drastic viral population bottleneck following the transition from rainy to dry season (**Fig. S4**). In the AM-Central/Northern regions, the inferred *Ne* remained relatively stable from August to October and increased during the rainy season from November to February (**Fig. S4**). These findings support that the OROV epidemic in the western Amazon Brazilian region resulted from synchronous sub-epidemics occurring in RR and AMACRO (2022-2023 rainy season) and in AMACRO and AM-Central/Northern regions (2023-2024 rainy season). Viral movements within each subpopulation were frequent, whereas migrations between them were sporadic. The viral demographic pattern coincides with the seasonal pattern of OROV+ cases and was probably driven by alternating periods of high viral transmission during rainy seasons and low, but persistent, viral transmission during dry seasons.

Analysis of genomic variations in the OROV_BR-2015-2024_ phylogeny reveals that several non-synonymous mutations were fixed across M (n = 11) and L (n = 7) genomic segments during recent evolution in the Amazon basin over the past decade (**Fig. 4e**), although most mutations fixed (90%) were synonymous. A total of 137 substitutions in the M (10 nonsynonymous and 47 synonymous), L (five nonsynonymous and 69 synonymous), and S (six synonymous) segments were detected at internal branches connecting the sequence sampled in Tefe-2015 with the MRCA of RR-I subclade, and 123 substitutions in the M (seven nonsynonymous and 34 synonymous), L (six nonsynonymous and 66 synonymous), and S (10 synonymous) segments at internal branches connecting Tefe-2015 with the MRCA of subclades circulating in AC, AM, and RO. Amino acid substitutions were mapped on glycoproteins, Gn (positions 1-300) and Gc (positions 482–1420), and the intervening NSm protein (positions 301-481) of the M segment. Interestingly, four substitutions occur in linear epitopes of Gn (8-28) and Gc (510-544, 616-632, and 706-718) glycoproteins recognized by the serum of immunized mice^46^ and two others occur in linear B-cell epitopes of Gc (1197–1208) and NSm (409-416) proteins predicted by the use of bioinformatics tools.^47^

The geographical coordinates of the patient’s residential area were employed to reconstruct the fine-scaled dispersion using a continuous spatial diffusion model with non-homogenous dispersion rates. The dispersion rate of viral lineages circulating in RR and AMACRO (RR-I+AMACRO-I+AMACRO-II) and those circulating in AM-Central (including Manaus) and AM-Northern regions (AM-I+AM-II+AM-III) were calculated separately (**Fig. 5a-b**). The average dispersion rate was estimated at 1.00 km/day (95% HPD: 0.79 - 1.26 km/day) in the RR/AMACRO regions and 0.66 km/day (95% HPD: 0.59 - 0.73 km/day) in the AM-Central/Northern regions, highlighting some differences in the OROV dynamics between these two areas (**Fig. 5c**). The viral dispersion rate also displayed some variation through time within each area (**Fig. 5d-e**). In the RR/AMACRO region, the viral dispersion rate remained stable at ∼1 km/day (0.75 - 1.23 km/day) from August 2022 to March 2023, but it progressively increased from May 2023, reaching 1.65 km/day (1.21 - 2.14 km/day) by the end of December 2023. In the AM-Central/Northern regions, the viral dispersion rate decreased slightly from 0.74 km/day (0.67 - 0.81 km/day) in August 2023 to 0.65 km/day (0.59 - 0.72 km/day) in February 2024. Thus, during the 2023-2024 rainy season, the rate of viral dispersion in the AMACRO region was ∼2.5x faster than in the AM- Central/Northern regions. Continuous phylogeographic reconstructions further revealed that very-short (<2 km), short (2–10 km), medium (11-30 km), and long (> 30 km) distance spread had a different impact on OROV dissemination, comprising 65%, 5%, 9% and 22% of all viral migrations, respectively (**Fig. 5f-g**).

**Figure 5.**
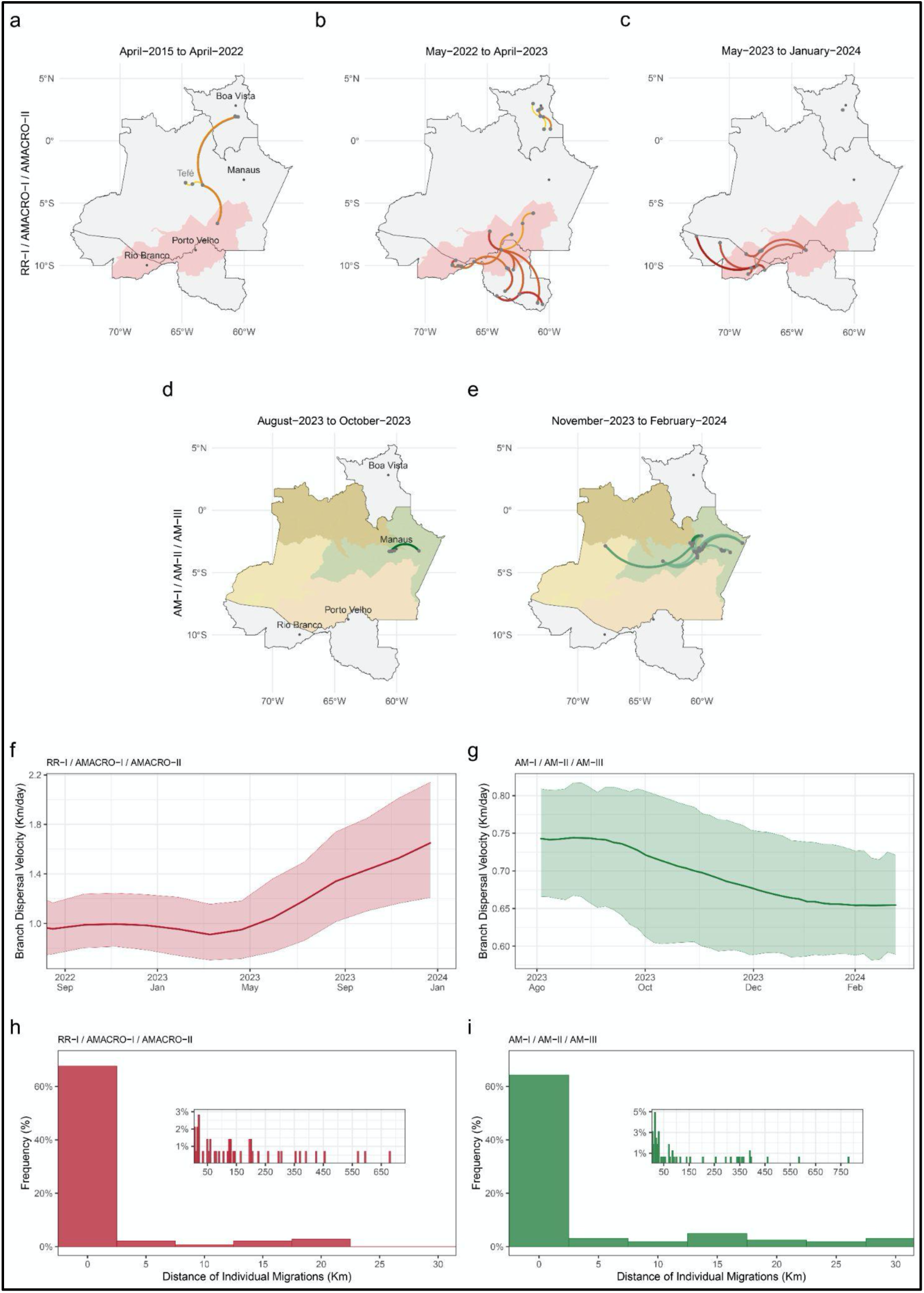
Spatiotemporal dispersal of the main OROV_BR-2015-2024_ sub-clades circulating in RR/AMACRO and AM-Central/Northern regions estimated from the continuous diffusion phylogeography process. **a-b)** Lines represent MCC tree branches of the RR/AMACRO (**a-c**) and AM central and northern regions subclades (**d-e**) projected on the maps according to the location of internal phylogenetic nodes and tips. Lines are shaded in gradients that transition from lighter to darker hues, representing the passage of time within each panel and connecting the origin and destination of migrations using a clockwise curvature. Major cities are annotated in the first panel of each sequence. **f-g)** OROV epidemic weighted branch dispersal velocity through time (posterior median = solid lines, 95% HPD = pale areas). **h-i)** Histograms of the frequency distribution of short (< 5km), medium (5 – 30 km), and long (> 30 km) distance viral migrations calculated from the branches of 1,000 randomly selected trees from the posterior distribution of the continuous phylogeographic analysis.

The pattern of OROV spread in the Amazon region, characterized by short-range (< 2 km) movements along continuous and contiguous areas with an average dispersal rate of ≤ 1 km/day, is entirely consistent with the pattern of active flight during the lifespan of infected vectors. Sylvatic mosquitoes in Brazil may move up to 3 km/day,^48^ and *Culicoides* spp. in North America display a mean dispersal rate of ∼2 km/day and typically travel <5 km from their breeding sites.^26,49,50^ Interestingly, the mean rate of spatial dispersal of OROV in the Amazon region is roughly similar to the spread rate of sylvatic transmission estimated for the YFV in the Americas (0.50 km/day)^51^ and Brazil (0.50-1.42 km/day).^52–54^ These findings suggest that the dispersal of infected vectors was probably a major driving force of the spatial spread of OROV in the Amazon basin. A significant proportion (31%) of OROV migrations, however, traveled medium and long distances (> 10 km) consistent with viral dispersion via infected humans. It is presumed that some individuals acquire the infection in the forest and later visit a rural or urban setting during the viremic phase, acting as a source of infection for biting midges and as the probable link between the sylvatic and urban transmission cycles of OROV.^1^ This potential link between sylvatic and urban transmission cycles of OROV adds a layer of complexity to disease control strategies. The expansion of transportation networks in the Amazon region also significantly increases human mobility and rapidly connects forest regions and rural settlements with urban areas, and may thus foster the spread of zoonotic infections among humans.^55^ The movement of infected individuals by terrestrial and air travel could have mainly contributed to the spread of OROV between non-contiguous rural and urban areas, rapidly expanding the geographic boundaries of viral transmission within the Amazon region and beyond.

While annual variations in rainfall may contribute to the seasonality of OROV transmission observed between 2023 and 2024, this factor does not explain the inter-annual variability in OROV epidemics nor the singular characteristics of the current OROV epidemic in the Amazon basin, which is distinguished from the previous ones by the longer duration and broader geographic extension. While previous outbreaks/epidemics generally last a single rainy season and impact one urban area or a few inner cities of a given state, the current epidemic spans at least two consecutive rainy seasons. The current outbreak affects over 140 municipalities across four northern Brazilian states, encompassing an area of approximately 2 million km^2^. Some additional key factors that operate on different timescales might have combined to alter the ecological dynamics of OROV in recent years, increasing the risk of zoonotic spillovers and the subsequent amplification of viral transmissions among humans in the Brazilian Amazon region. Viral evolution, anthropogenic landscape perturbations, and global climate changes are critical drivers of oscillations in the incidence and geographic spread of arboviral diseases in human populations.^56^

OROV evolution can occur in very short timescales due to the high mutation rate and the capacity of genetic reassortment, allowing the virus to acquire key adaptive mutations rapidly.^57^ We hypothesize that the upsurge of OROV cases in western Amazon was driven by the emergence of a novel reassortant clade OROV_BR-2015-2024_ with enhanced viral transmissibility in sylvatic and/or urban cycles. However, the clade OROV_BR-2015-2024_ circulates endemically in the Amazon basin for almost a decade without causing large outbreaks.^31^ It could be argued that clade OROV_BR-2015-2024_ accumulated key mutations since 2020 that potentially increased viral transmissibility. However, the lack of similar large OROV epidemics in the eastern Brazilian Amazon region (PA and AP states) does not support this hypothesis. Further studies are essential to compare the replicative and infective potential between past and current circulating OROV lineages and to test if some mutations fixed during the recent evolution of OROV may impact virological traits such as infectivity or immune evasion. Without these data, we cannot rule out that additional factors beyond viral evolution drive the explosive OROV epidemics in the western Amazon between 2022 and 2024.

Extreme environmental changes may influence the long-term transmission dynamics of arboviruses,^58–61^ and current OROV outbreaks in the Amazon region coincide with remarkable El Niño Southern Oscillation (ENSO) events and extreme climatic events. The period of silent dissemination of the novel OROV reassortant strain (2011-2021) coincides with a high frequency of severe floods in the Amazon basin.^62–64^ The current OROV epidemic, from November 2022 to March 2024, was foregone by unprecedented flooding events in the southwestern Amazon region during 2020-2021 caused by a rare multi-year 2020-23 La-Niña event.^65,66^ Moreover, the current OROV epidemic overlapped with the transition from the 2022-23 La Niña to the 2023-24 El Niño, which caused record drought and heat in the Amazon basin.^67^ Southwestern Amazonia experienced negative rainfall anomalies from November 2022 to February 2023, while northern Amazonia faced damaging rainfall and record-high temperatures from June to September 2023. Climatic extremes have become more widespread and frequent across Amazon^68^, which may have changed the OROV endemic and epidemic transmission dynamics.

Fragmented forest landscapes and vegetation loss due to deforestation and expanding agricultural land use were pointed as key drivers of OROV transmission.^69–71^ Alarming growing deforestation trends have been observed in the Brazilian Amazon region since 2018, mainly driven by illegal land grabbing, timber extraction, mining, and agricultural expansion into forested lands.^72^ Most OROV-positive localities in 2022-2023 were concentrated in AMACRO, an area considered the new frontier of deforestation that was responsible for a significant fraction of forest lost in the AM (82%), RO (77%), and AC (63%) states between 2017 and 2021.^72–74^ Similarly, critical deforestation areas in the RR state were close to the municipalities of Alto Alegre, Mucajaí, Iracema, and Rorainópolis,^72^ which concentrated most OROV-positive cases in the state. The weakening of environmental governance in Brazil between 2018 and 2022 certainly acted as a stimulus for the advance in deforestation across all Amazonian regions.^75–78^ The accelerated deforestation in the AMACRO region further coincided with its designation as a special zone for agricultural development,^79–81^ the expansion of agribusiness activities reliant on expanding frontiers by clearcutting forests,^79,82^ and the improvement of the infrastructure like the highway BR-319, which connect the state capitals of AM and RO and crosses several conservation units and Indigenous lands.^75,83–86^

Although establishing direct causality between events is challenging, the circumstantial evidence described above points to a putative link between current OROV outbreaks and extrinsic environmental factors that culminate in higher pathogen exposure to the human population in Brazilian Amazon.^87^ Synergistic interactions between uncontrolled deforestation, agricultural encroachment, road construction, extreme flows, and warming droughts over the last few years may have altered the ecological conditions for endemic/urban OROV transmission and further increased the number of susceptible people near new forest edges in the western Brazilian Amazon. These factors combined increase the risk of viral spillovers from wildlife to humans and the subsequent human-to-human transmissions within rural and urban settings of the Amazon region. The absence of similar OROV outbreaks at deforestation frontiers in the eastern Amazon (AP and PA states) might be explained by intrinsic factors such as a lower abundance of susceptible hosts. Due to the longer history of forest degradation and OROV outbreaks, people from the eastern Amazon could have been more frequently exposed to the virus, making them less susceptible to infection than people from neo-colonized areas in the western Amazon.

Our study has some limitations. First, the 7-year gap between the earliest sampled OROV sequence of the current outbreak and the most closely related non-outbreak sequence sampled in Tefe in 2015 limits our ability to reconstruct the pattern of viral spread pre-2022 accurately. Second, geographic-based sampling bias during the current outbreak may have affected the accuracy of phylogeographic reconstructions. Hence, many apparent long-distance viral migrations may have resulted from sequential shorter-range migrations between unsampled locations.^88–90^ Despite this potential limitation, phylogeographic inferences performed using different datasets (complete or downsampled) and methods (discrete or continuous) recovered similar spatiotemporal patterns. Third, the absence of long-term laboratory surveillance of OROV cases in Brazil hinders our ability to determine whether recent upsurges are due to enhanced surveillance efforts or to enhanced viral transmission. Finally, we have no information about the primary vector involved nor about temporal changes in vector abundance and biodiversity across sylvatic, forest edges and urban environments in more affected areas during the ongoing 2022-2024 OROV outbreak.

In summary, our findings revealed that a new reassortant OROV lineage that circulated in the Amazon region for about one decade caused an upsurge of OROV cases in several rural and urban settings of the western Amazon Brazilian region between 2022 and 2024. Our results confirm the high epidemic potential of OROV in the Amazonian biome and raise concerns about the risk of spreading and establishing this neglected arbovirus outside the Amazon region. Indeed, recent findings support that the OROV_BR-2022-2024_ clade was successfully established in the Southeastern and Southern Brazilian regions and the Caribbean region by early 2024.^21^ Our study also warns about the crucial need for the widespread distribution of diagnostic tests for the detection of OROV across all Brazilian states. Further studies will be needed to assess whether the recent evolution of OROV might have changed vector competence and virus transmissibility in wild or urban cycles. Moreover, the implementation of long-term interdisciplinary "One Health" disease surveillance systems is needed to understand the relationship between extrinsic environmental factors, wildlife biodiversity, and the dynamics of OROV infections in human and wild reservoir hosts. Such information is crucial to identify “pathogenic landscapes” that may increase the risk of OROV transmission^91^ and to develop early warning systems for OROV epidemics.

## METHODS

### OROV positive samples and ethical aspects

Acute OROV infection was detected using a duplex reverse-transcription real-time PCR assay previously developed by our team in Manaus that simultaneously detects OROV and Mayaro viruses.^22^ This protocol was recently implemented in all Central Public Health Laboratories located in each Brazilian State (LACEN) by the Brazilian Ministry of Health and was also recommended by the Pan-American Health Organization for the molecular diagnosis of OROV and Mayaro viruses (https://www.paho.org/en/documents/real-time-rt-pcr-protocol-mayaro-mayv-and-oropouche-virus-orov-duplex). The Central Laboratories from the States of AM (LACEN-AM), RR (LACEN-RR), RO (LACEN-RO), and AC (LACEN-AC), as well as Fiocruz Rondônia, sent OROV positive samples for sequencing at Instituto Leônidas e Maria Deane (ILMD - FIOCRUZ Amazônia), that is the hub of FIOCRUZ Genomics Surveillance Network of the Brazilian Ministry of Health in Manaus, AM for viral genome characterization. Information regarding the week and location of the collected samples analyzed in this study and their phylogenetic context (considering the M segment) may be visualized in detail on our Microreact ^39^ page: https://microreact.org/project/orov-north-2022-2024. This study was approved by the Ethics Committee of Amazonas State University (CAAE: 72678923.8.0000.5016), which waived signed informed consent.

### Epidemiological data

#### Individual-level data

case line-list data was retrieved from GAL (Gerenciador de Ambiente Laboratorial) at the municipality level for the states of Acre, Amazonas, Roraima, and Rondônia between January 2022 and March 2024. For Amazonas, all arboviral notifications for the period were recorded. Each case had a notification code and metadata associated with it which included demographic characteristics, spatial location, laboratory results and clinical details such as symptoms, hospitalization status, and dates associated with notification, and onset with these were reported at least for most cases. For the other states, only the information about OROV detection was retrieved. Identifiers were anonymized. <u>Data Cleaning:</u> We identified and merged any duplicate entries in the line-list. And reconciled dates where onset date was in discrepancy with notification date. <u>Demographic data.</u> Demographic data for ages at the level of municipality from the 2022 census was retrieved from the Brazilian Institute of Geography and Statistics (https://sidra.ibge.gov.br/pesquisa/censo-demografico/demografico-2022/universo-populacao-por-idade-e-sexo). These data were used to calculate age specific incidence of reported OROV cases. <u>Medical records:</u> An investigation of the clinical symptoms was conducted by the Amazonas State Health Surveillance Foundation - Dr Rosemary Costa Pinto (FVS-RCP) among the 2,272 OROV-positive patients from the AM state who answered the medical form in 2024.

### OROV amplification and sequencing

We sequence one full-length OROV genome (Tefé-2015) using an unbiased metatranscriptomics protocol (Illumina Ribo-ZeroTM Plus Microbiome Depletion Kit). This comprehensive sequencing, along with other complete genomes available in GenBank, served as a pivotal reference for the design of an amplicon-based strategy using Primal Scheme ^92^ and a modified version of Primer3 ^93^ embedded in Geneious Prime 2023.2.1 (**Supplementary Table 2**). All other OROV genome amplification and sequencing libraries were prepared with Illumina COVIDSeq backbone for a more cost-effective genome-wide sequencing strategy. However, with our newly OROV-designed primers on 2 × 150 cycles paired-end runs. Raw sequencing data was collected with MiSeq Control Software v2.6.2.1 or NextSeq 1000 Control Software Suite v1.2.0. Subsequently, it was converted to FASTQ files on Illumina’s cloud-based application (https://basespace.illumina.com). FASTQ files were imported to Geneious Prime v2023.2.1, trimmed, and assembled into contigs using a workflow customized by our group employing BBDuk, Dedupe, and BBMap tools (v.38.84), embedded in Geneious Prime. The GenBank sequences OL689332, OL689333, and OL689334 were used as templates for the S, M, and L Oropouche virus segments. All contigs were visually inspected before consensus calling using a threshold of at least 50%. Finally, using this sequencing strategy, we were able to recover 87.2%, 98.8%, and 98.3% of the S, M, and L segments, respectively.

### OROV whole-genome consensus sequences and genotyping

The 383 complete OROV consensus sequences of S, M, and L genomic segments generated in this study (382 from the current outbreak plus one from Tefé-AM 2015) were aligned with corresponding segments of all published full-length OROV genome sequences available at the NCBI (n = 72), sampled in the Americas between 1955 and 2021, including the prototype sequences of OROV (OROV_P_ L: AF484424; M: AF441119; S: AY237111), Iquitos virus (IQTV_P_ L: KF697142; M: KF697143; S: KF697144), Perdoes virus (PEDV_P_ L: KP691627; M: KP691628; S: KP691629), and Madre de Dios virus (MDDV_P_ L: KF697147; M: KF697145; S: KF697146). All these viruses are classified in the species *Orthobunyavirus oropoucheense*, having the classical OROV as the exemplar isolate of the species (https://ictv.global/report/chapter/peribunyaviridae/peribunyaviridae/orthobunyavirus). The resulting datasets were used to infer Bayesian phylogenetic trees for each genomic segment.

Bayesian phylogenetic trees were reconstructed under the best-fitted substitution model selected by jModelTest 2.1.10^94^, using the MrBayes 3.2.7a program ^95^. Two chains were run for 20 × 10^6^ generations for each genomic segment, and the attainment of the stationary phase and the effective mixing of continuous parameters (effective sample size [ESS] > 200) were assessed using Tracer v1.7^96^. The consensus trees were visualized using the Treeio v3.1.7^97^ and ggtree v3.2.1 R packages^98^. Clusters were defined by their statistical support (*posterior probability* [*PP*] > 0.90).

### Bayesian evolutionary, phylogeographic, and demographic analyses

The temporal structure of the selected sequences was assessed using the program TempEst v.1.5.3^99^ by a root-to-tip linear regression, in which the significance of the correlation between the collection date and genetic distance was accessed with a Spearman correlation test. Time-scaled phylogenetic trees for separate or concatenated genomic segments were estimated using the Bayesian Markov chain Monte Carlo (MCMC) approach, implemented in the software BEAST 1.10^100^ with BEAGLE library v.3^101^, to improve computational time. Bayesian trees were reconstructed using the GTR+G4+I nucleotide substitution model, the non-parametric Bayesian skyline coalescent demographic model,^102^ and a relaxed molecular clock model with a continuous-time Markov chain (CTMC) rate reference prior.^103^ We also explicitly modeled the OROV reassortment process in a Bayesian framework by applying the Coalescent With Reassortment Constant Population (CoalRe) model as available in the BEAST2 package.^45^ An exponential distribution with a mean of 0.25 was set as the prior on the reassortment rate parameter. The analysis parameterization was complemented with the GTR+G4+I substitution model and a relaxed clock with a lognormal distribution model with default priors. As explained above, the spatial dispersion pattern of OROV in the Amazon region was reconstructed using both discrete and continuous phylogeographic analyses implemented in BEAST v.1.10.4.^100^ For the discrete phylogeographic analysis, we employed a reversible discrete phylogeographic model^104^ with a continuous-time Markov chain (CTMC) rate reference prior.^103^ OROV sequences from AC, RO, and RR were grouped by state. In contrast, sequences from the state of AM were grouped by sub-state regions (AM-Central, AM-Southern, AM-Northern and AM-Southwestern), with the only exception of sequences from the capital city, Manaus, which belongs to the AM-Central region, but were grouped in a separate location due to the significant contribution of this city to the overall number of OROV cases and sequences. For the continuous phylogeographic reconstruction, we employed the Cauchy relaxed random walk model.^105^ For the continuous phylogeographic analyses, a maximum of 10 genomes per municipality (the earliest ones) were chosen to reduce the impact of within-municipality transmissions on the estimation of viral diffusion rates among Amazon municipalities. We assigned latitude and longitude coordinates to each sequence according to the municipality of origin. We selected the option *Add random jitter to tips*, adding noise to sampling coordinates to ensure unique geographic coordinates for each sequence. Multiple independent Markov chain Monte Carlo (MCMC) runs were performed and later combined to ensure that all continuous parameters had an effective sample size (ESS) > 200, as visualized in Tracer v1.7.^96^ The maximum clade credibility (MCC) trees were summarized with TreeAnnotator v.1.10 and visualized using FigTree v.1.4.4 (http://tree.bio.ed.ac.uk/software/figtree/). Reassortment networks were visualized and edited in IcyTree.^106^ Viral spatiotemporal diffusion was analyzed and visualized in SPREAD v.1.0.7^107^ and further projected in maps generated with the ggplot2,^108^ sf,^109^ and geobr^110^ R packages.

### Statistical analyses

The Fisher exact test (two-tailed) was used to compare the frequency of symptoms between males and females infected with OROV. The threshold for statistical significance was set to *P* < 0.05 using two-sided tests. Statistical analyses were performed using GraphPad v.9.0 (Prism Software).

## Data availability

All the OROV genomes generated and analyzed in this study were deposited at GenBank under accession numbers PP153945 to PP154172 and PQ064571 to PQ065491.

## Acknowledgments

The authors wish to thank all the healthcare workers from the Brazilian States fighting the current Oropouche outbreak. We also appreciate the support of FIOCRUZ COVID-19 Genomics Surveillance Network members, the General Laboratory Coordination (CGLab) of the Brazilian Ministry of Health (MoH), Brazilian Central Laboratory States (LACENs) and health surveillance agencies from Acre, Amazonas, Rondônia, and Roraima. Funding support FAPEAM Call 04/2022/FIOCRUZ/FAPEAM/FAPERO - INOVAÇÃO NA AMAZÔNIA; Amazônia +10; FAPEAM Call 023/2022 - INICIATIVA AMAZÔNIA +10; Inova Fiocruz - Inova Amazônia; Rede Genômica de Vigilância em Saúde do Estado do Amazonas – REGESAM (Resolução n°. 002/2008, 007/2018 e 005/2019 – PRÓ-ESTADO/FAPEAM); Conselho Nacional de Desenvolvimento Científico e Tecnológico - CNPq Institutos Nacionais de Ciência e Tecnologia (INCT - VER) e Chamada CNPq/MCTI 10/2023 - Faixa B - Grupos Consolidados - Universal 2023 (421620/2023-4). GLW and GB are supported by the Conselho Nacional de Desenvolvimento Científico e Tecnológico (CNPq) through their productivity research fellowships (307209/2023-7 and 304883/2020-4). ED and AR are supported by Fundação de Amparo à Pesquisa e Inovação do Espírito Santo (FAPES).

---

**Supplementary Table 1.** Signs and symptoms observed in Oropouche fever patients from Amazonas State, Brazil, 2024.

| **Signs/Symptoms** | **Female**<br>**N = 1,085** | **Male**<br>**N = 1,187** | **Two-tailed**<br>**Fisher exact test** |
| --- | --- | --- | --- |
| Fever | 1,044 (96.8) | 1,153 (97.5) | 0.2408 |
| Headache | 1,012 (93.8) | 1,084 (91.7) | 0.0845 |
| Myalgia | 854 (79.1) | 913 (77.2) | 0.3127 |
| Nausea | 501 (46.4) | 450 (38.1) | **> 0.0001** |
| Retro-orbital pain | 367 (34) | 403 (34.1) | 0.9646 |
| Arthralgia | 313 (29) | 332 (28.1) | 0.6751 |
| Vomiting | 314 (29.1) | 268 (22.7) | **0.0005** |
| Photophobia | 101 (9.4) | 89 (7.5) | 0.1293 |
| Diarrhea | 89 (8.2) | 79 (6.7) | 0.1726 |
| Rash | 35 (3.2) | 28 (2.4) | 0.2496 |
| Edema | 35 (3.2) | 28 (2.4) | 0.2496 |
| Other<br>signs/symptoms | 330 (30.6) | 361 (30.5) | 1.0000 |

In 2024, 2,272 Oropouche fever patients from the Amazonas State, 1,085 females, and 1,187 males, answered the medical form. The absolute number for each sign/symptom and the percentage (in brackets) related to the total number for each gender are shown. The Two-tailed Fisher exact test results are shown in the fourth column, and those statistically significant are highlighted in bold.

**Supplementary Table 2.**
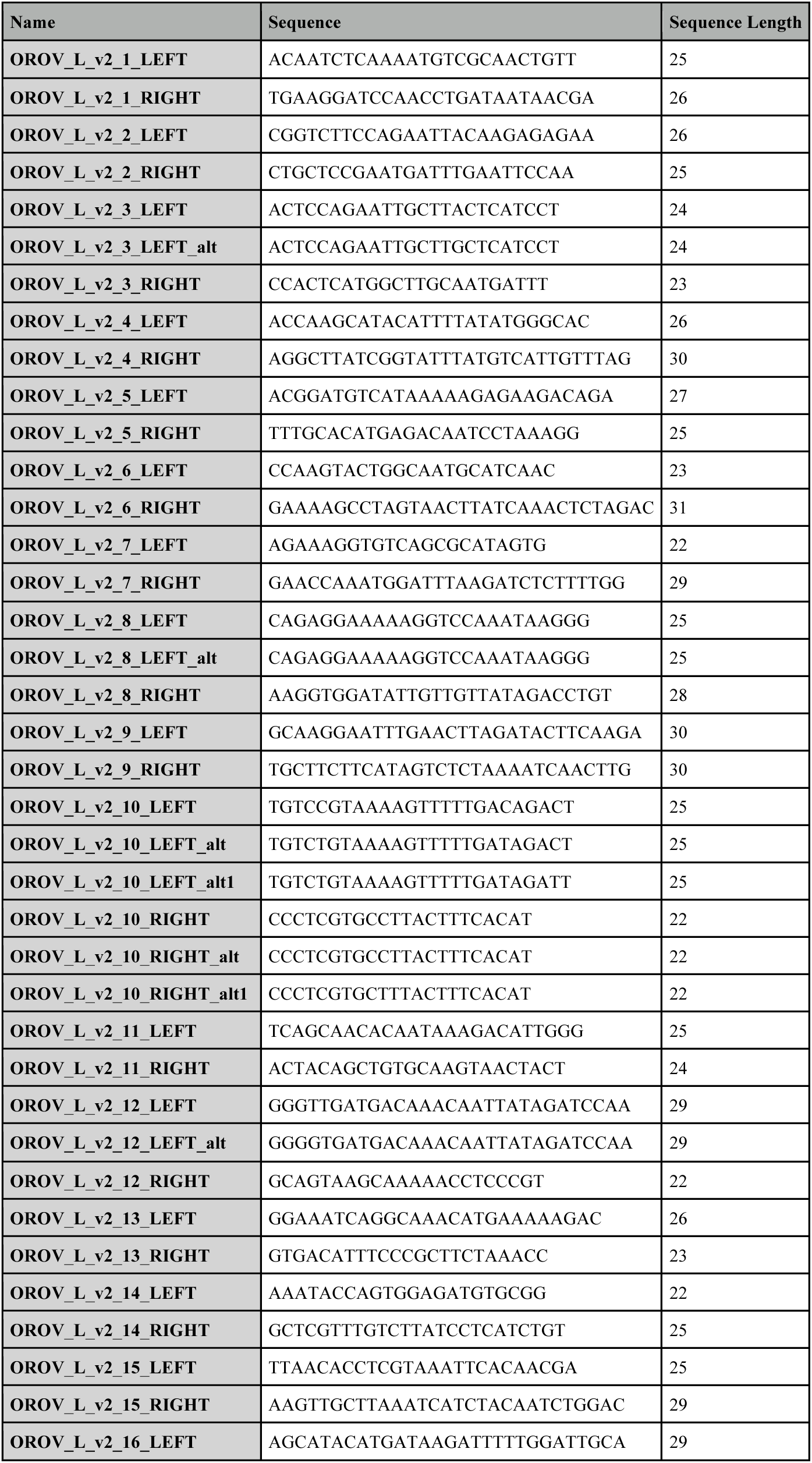
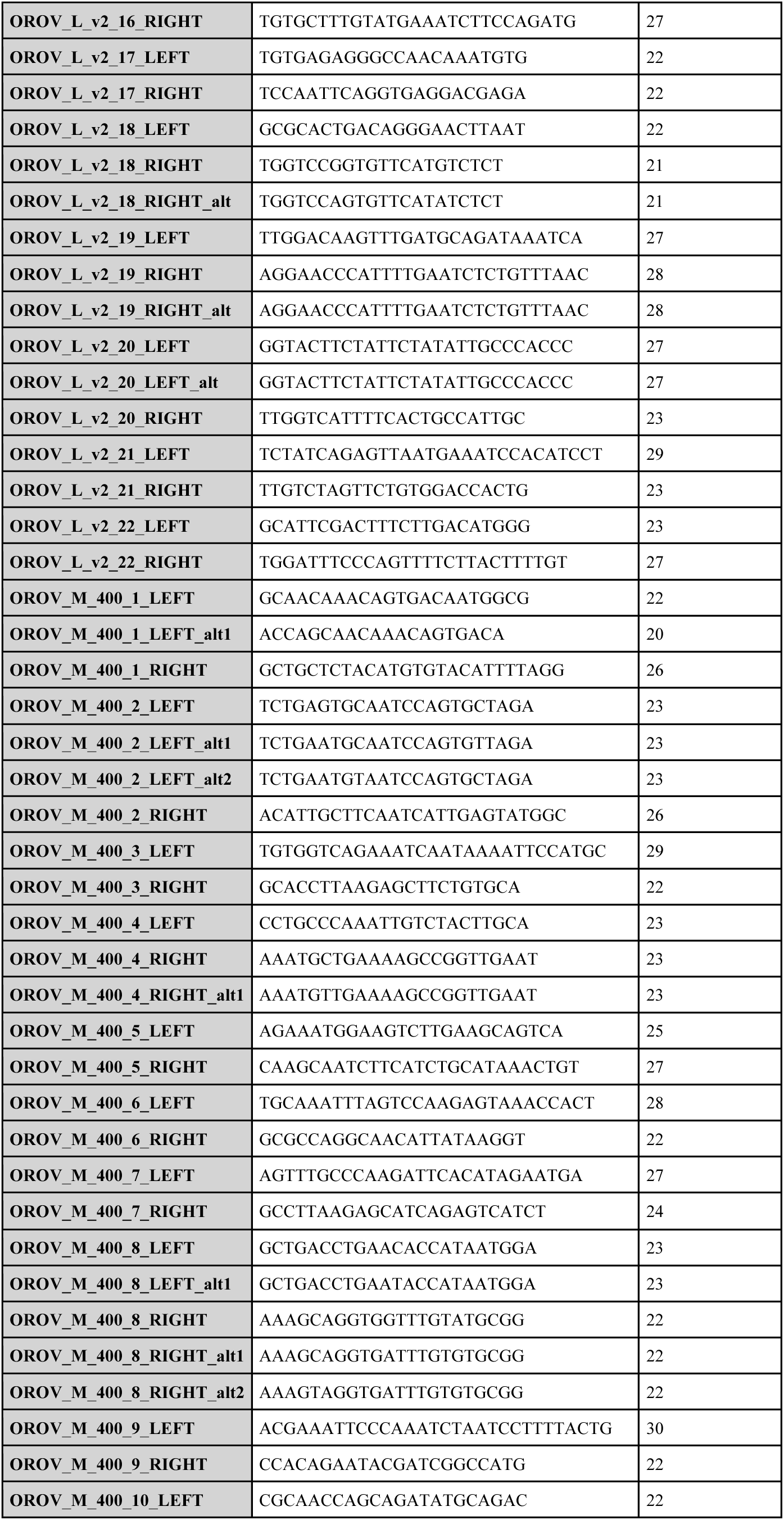
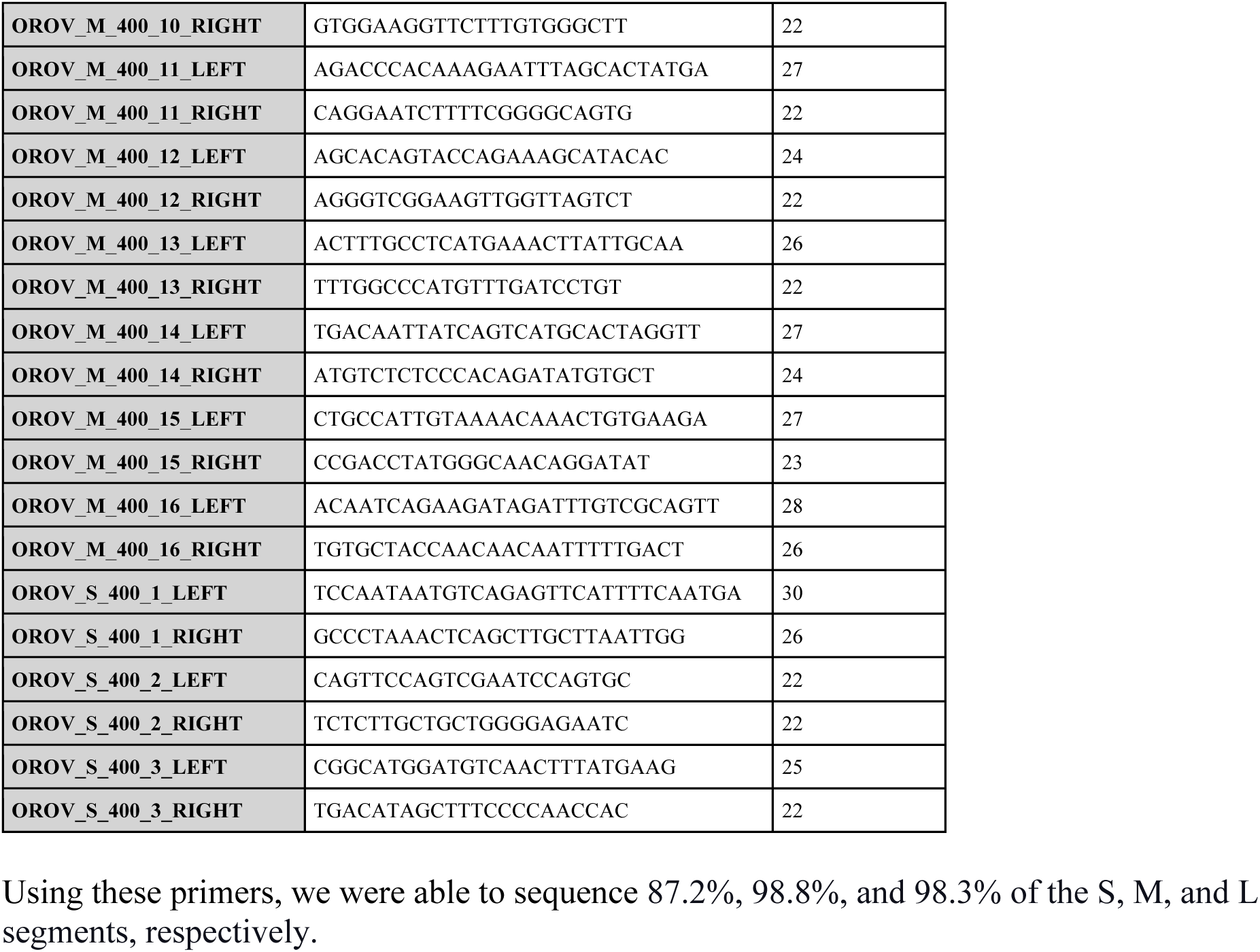
Primers designed and used to sequence near-to-complete Oropouche virus genomes in this study.

**Figure S1.**
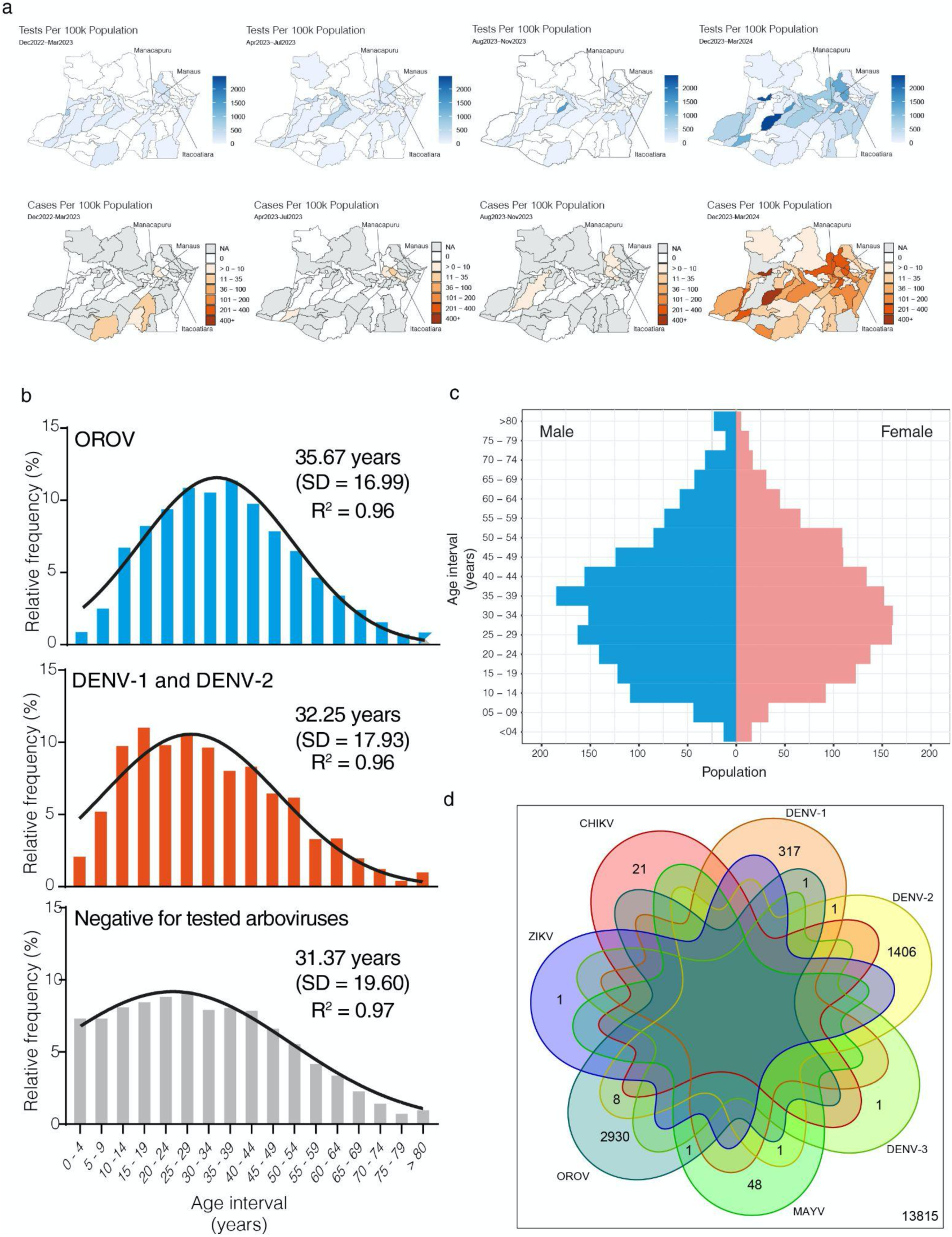
Comprehensive analysis of arboviral infections in the Amazonas (AM) state (2022-2024). a) Geographic distribution of RT-PCR tests for arboviruses (top panel) and OROV-positive cases per 100,000 population (bottom panel) in AM across four time periods: December 2022 to March 2023, April 2023 to July 2023, August 2023 to November 2023, and December 2023 to March 2024. Major cities (pop > 100k hab.) are indicated. B) Histograms depicting the age distribution of study patients in AM with different arboviral diagnosis profiles. Samples classified as ARBO-negative tested negative for OROV, Dengue virus (DENV), Zika virus (ZIKV), Chikungunya virus (CHIKV), and Mayaro virus (MAYV). C) Age-sex pyramid of the OROV-positive population from AM from 2022 to 2024, with males depicted on the left and females on the right. D) Venn diagram showing the distribution of detected viruses in the analyzed samples and the co-infections.

**Figure S2.**
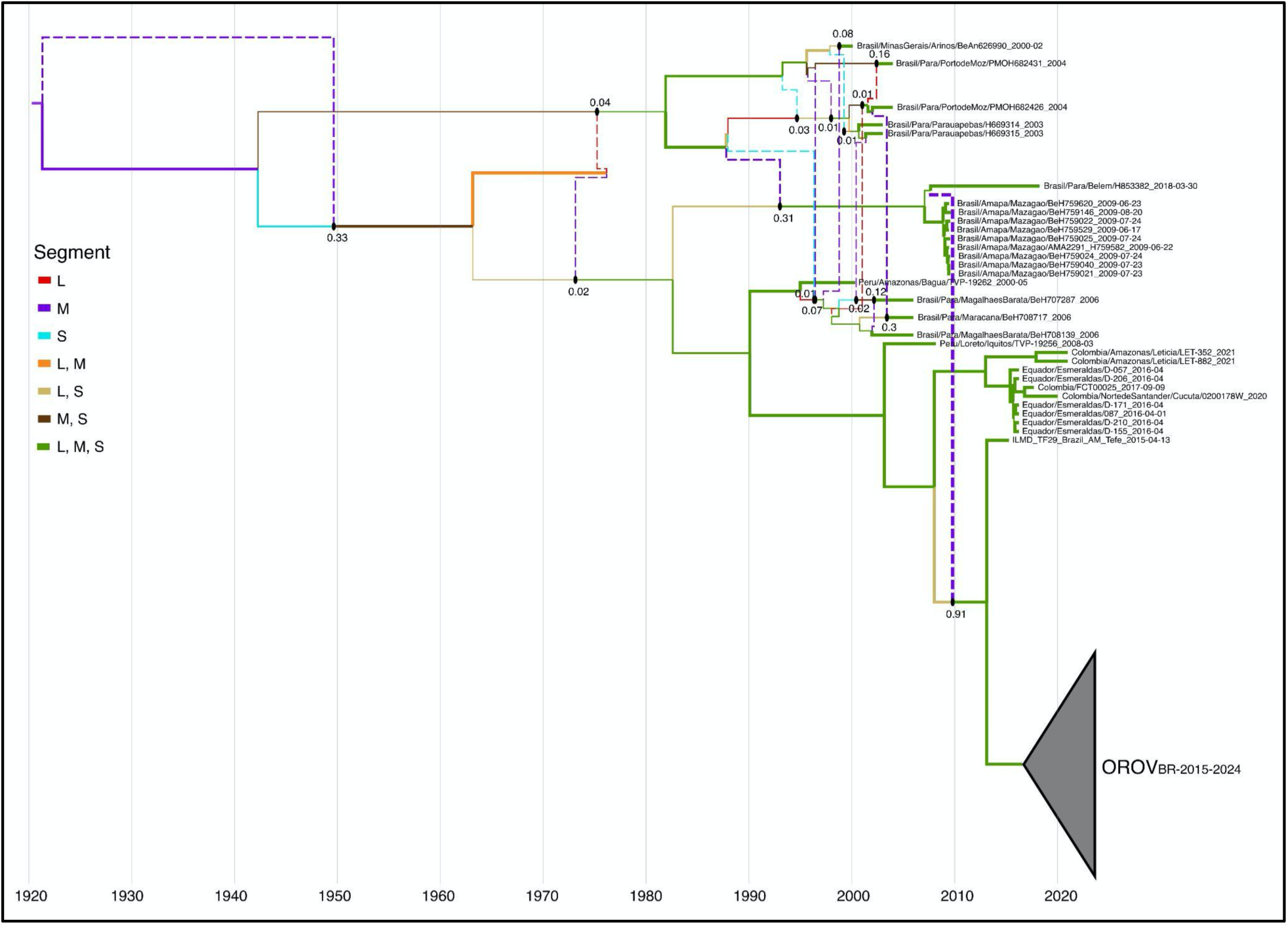
Maximum clade credibility (MCC) network of OROV complete genomes sampled after 2000. Reassortment events are represented as dashed-lines and the corresponding *PP* is shown. Branches thickness are according to the branch pp and colors represent the OROV segments carried by the branch as in the legend.

**Figure S3.**
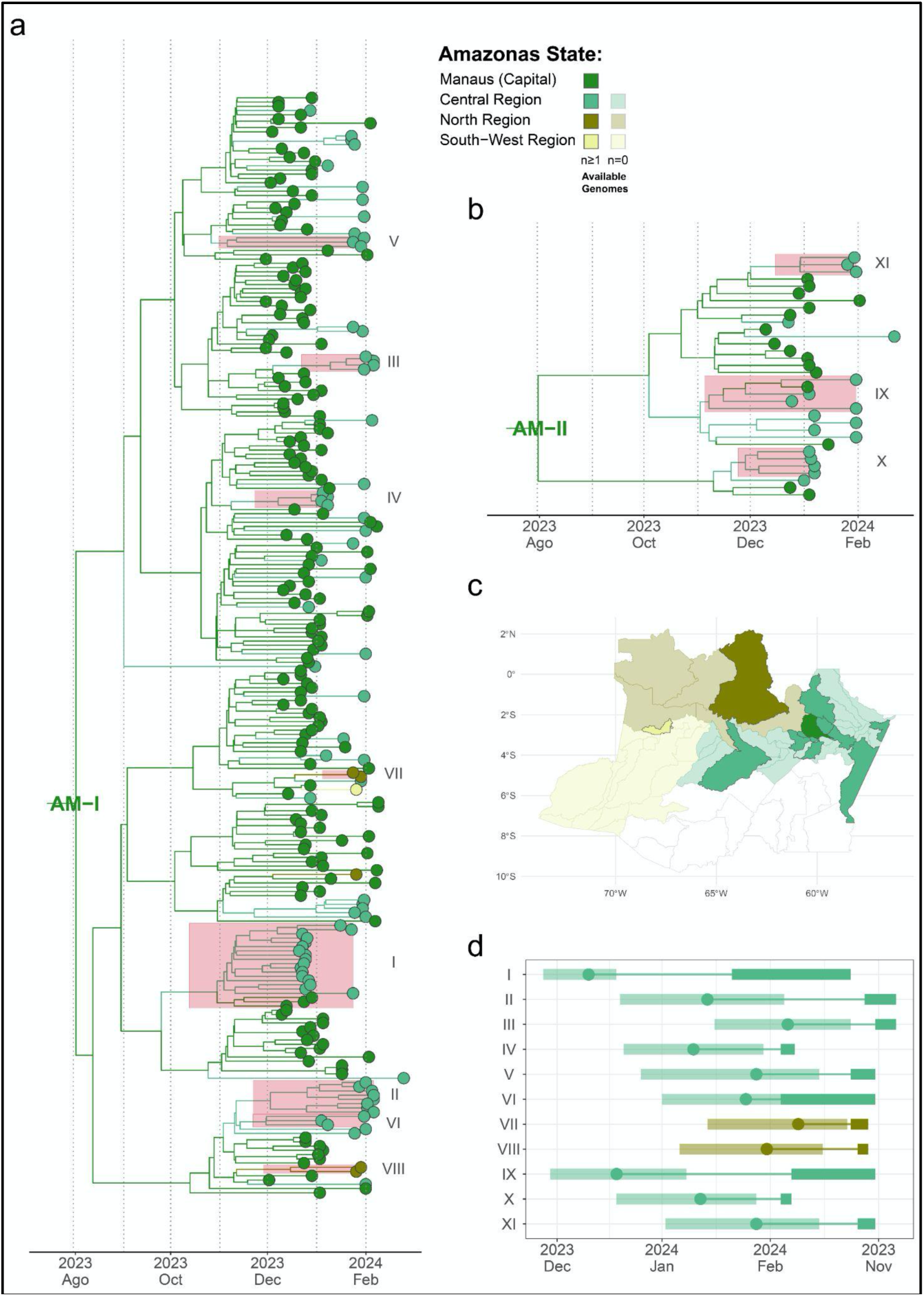
Spatial dissemination of the OROV_BR-2015-2024_ clade in AM-Central/Northern regions. **a-b**) Time-scaled MCC tree of the OROV_BR-2015-2024_ AM-I (*n* = 259) (**a**) and AM-II (*n* = 34) (**b**) sub-clades, inferred after concatenation of the three genomic segments (L, M, and S) of each sample. Branches are colored according to the inferred location of their ancestor nodes, and tips are colored according to sampling locations, both following the color scale shown in the top right corner of the AM-I tree. Sub-clades are highlighted in the tree alongside their posterior probabilities. **c**) Map of Amazonas state, showing its sub-state regions, according to the same color scale used in the MCC tree. In each region, cities with available genomes are colored in a darker hue, as indicated in the legend. **d**) Temporal dynamics of major AM-I and AM-II sub-clades. For each sub-clade, we represent the time of the most recent common ancestor (T_MRCA_, *circle*) and its 95% highest posterior density (HPD) interval (*transparent polygon*), the period of cryptic circulation (*thinner line*), and the sampling range (*thicker line*). Each sub-clade is colored following the same color scale used in the MCC tree.

**Figure S4.**
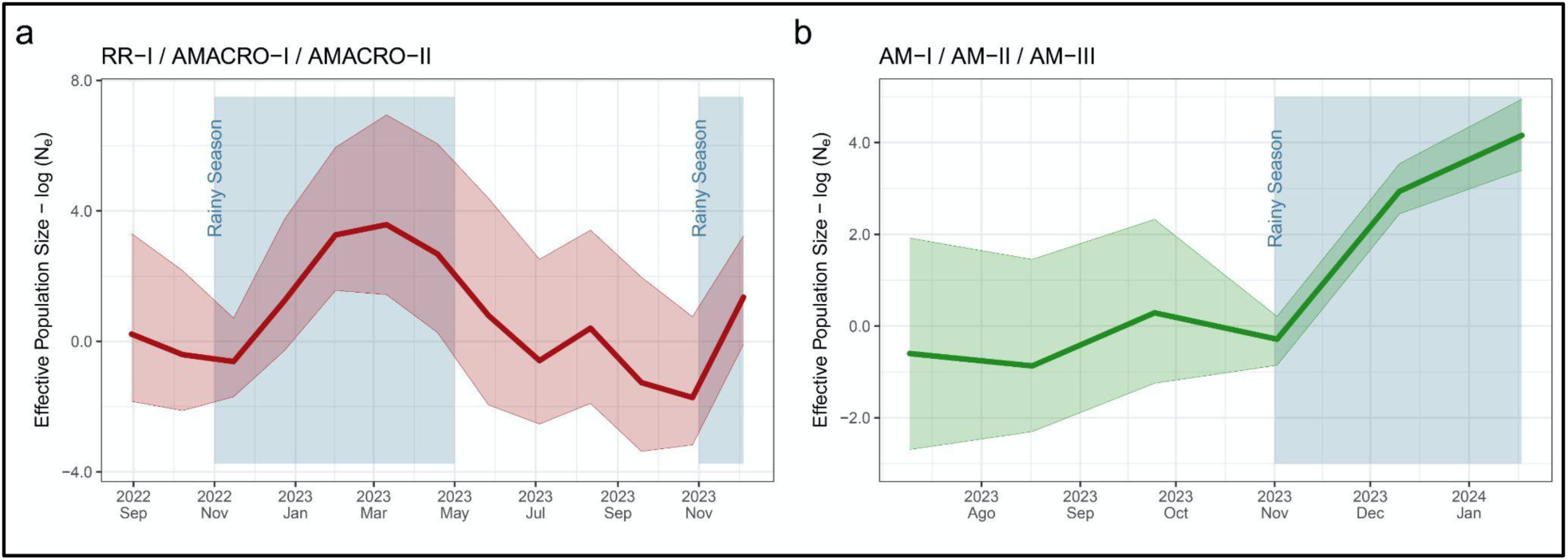
Demographic history of the main OROV_BR-2015-2024_ sub-clades circulating in RR/AMACRO (a) and AM-Central/Northern (b) regions. Each plot details the effective number of OROV infections (*Ne*, y-axis) over time estimated under the coalescent-based Bayesian Skygrid (BSKG) model (posterior median = solid lines, 95% HPD = pale areas). Blue-shaded areas indicate the Amazon basin region’s rainy season (November through May).

